# *Anopheles* salivary antibody biomarkers as surrogate outcomes measures to assess the effectiveness of topical repellent in Southeast Myanmar

**DOI:** 10.64898/2026.03.02.26347466

**Authors:** Ellen A Kearney, Paul A Agius, Katherine O’Flaherty, Win Han Oo, Julia C Cutts, Win Htike, Daniela Da Silva Gonçalves, Aung Thi, Kyaw Zayar Aung, Htin Kyaw Thu, Myat Mon Thein, Nyi Nyi Zaw, Wai Yan Min Htay, Aung Paing Soe, Julie A Simpson, Freya JI Fowkes

## Abstract

**Background:** Measurement of human antibodies against *Anopheles* salivary proteins that are injected during mosquito bites may serve as biomarkers of mosquito biting exposure. These biomarkers have been suggested as surrogate outcomes in trials of vector control intervention effectiveness, however, studies to-date have been largely descriptive and do not directly quantify the instantaneous nor cumulative effects of the intervention on antibody outcomes. In this study, we sought to explore the use of a serological biomarker of *Anopheles* spp. biting exposure as a surrogate outcome in a trial of topical repellent in Southeast Myanmar.

**Methods:** In a stepped-wedge cluster randomised controlled trial of personal repellent delivered to 114 villages, we measured the anti-gSG6-P1 IgG antibody response in 14,128 samples by high-throughput ELISA. Generalised linear mixed modelling was used to estimate the effects of repellent distribution (rolled out monthly to blocks of villages) on antibody outcomes overall and for high-risk populations (migrants and forest goers).

**Results:** Overall, we detected generally high levels (median: 2.1OD) and seroprevalence (59.9%) of anti-gSG6-P1 IgG antibodies. We observed no instantaneous effect of repellent on antibody levels to *Anopheles* salivary proteins (mean difference (MD)= 0.01; 95%CI=−0.03, 0.05), however estimation of a series of lagged effects of repellent distribution (*i*.*e*. modelling a gradual antibody decay from prolonged use) showed reduced antibody levels after transition to repellent (*i*.*e*. repellent distribution 6-months prior saw a 0.03-unit (95%CI= −0.08, 0.03) decrease in antibody levels). More specifically, we observed reductions in antibody levels for migrants (6-month lag: MD= −0.10; 95%CI= −0.21, −0.01) and forest dwellers (MD= −0.05; 95%CI=−0.10, 0.00), but not village residents (MD= 0.02; 95%CI=−0.04, 0.08).

**Conclusions:** These findings suggest antibodies to *Anopheles* salivary proteins could be an informative trial outcome measure and provide important parameters on antibody longevity to inform the design of future studies assessing the effectiveness of vector-control interventions.

## Background

Personal protection interventions may have important application in the Greater Mekong Subregion (GMS) to target residual malaria transmission and high-risk populations to accelerate progress towards elimination. However, the World Health Organization currently makes conditional recommendations against the use of supplementary personal protective interventions including topical repellents (1), citing a lack of robust evidence for their protective effect (2). Field trials of personal protective interventions effectiveness typically target parasitological rather than entomological endpoints; evidenced by the inclusion of only one study (3) in a 2023 Cochrane systematic review of topical repellents that estimated the effect of repellent on a traditional entomological indicator: human biting rate (HBR) (2). Defined as the number of bites a person receives per unit-time (and essential to measure the entomological inoculation rate [EIR: the gold standard measure of malaria transmission intensity]); measurement of HBR relies upon direct mosquito capture typically through methods like the “gold standard”: human landing catch. These approaches require volunteers to capture mosquitoes that land on their exposed limbs, and are therefore resource- and labour-intensive, and largely infeasible to conduct in field trials to measure changes in individual level exposure. Typically, estimates of HBR and EIR are aggregated across collectors to produce estimates of exposure at a population-level only (4, 5) which precludes exploration of heterogeneity due to individual-level factors that may influence a person’s exposure to vector bites (*i*.*e*. migrancy status, occupational exposure, etc.). Therefore, new and innovative tools that can measure changes in mosquito biting exposure at both an individual and population level are needed to assess the effectiveness of vector control interventions. This would have particular application in trials of interventions of personal protection targeted at preventing residual transmission and high-risk populations (*e*.*g*. mobile and migrant populations, those dwelling/working in forests, etc.), especially in elimination settings where low infection rates may impact on the statistical power of studies to detect small changes in the outcome (5, 6).

Human antibodies against *Anopheles* salivary proteins have been investigated as a logistically feasible alternative approach to quantify *Anopheles* biting exposure (7, 8). A 2021 systematic review identified that IgG antibodies against the most commonly investigated *Anopheles* salivary antigen, the *An. gambiae* Salivary Gland 6 (gSG6) protein, were associated with *Anopheles* spp. human biting rate (9), suggesting the potential use of these antibodies as a surrogate outcome measure in vector control intervention effectiveness trials (particularly for personal protection). This measure would be particularly relevant in low transmission settings where low numbers of events significantly impact the statistical power of the study. However, evidence for the utility of this surrogate measure is scant, as the majority of the vector control intervention effectiveness studies that measure antibodies specific for *Anopheles* salivary antigens are descriptive only and do not directly quantify the effect of the intervention on antibody outcomes (10–12).

In the present study, we seek to explore the use of a serological biomarker of *Anopheles* biting exposure as a surrogate outcome in a stepped-wedge cluster randomised controlled trial of topical mosquito repellent delivered by village health volunteers (VHVs) to 114 villages in Southeast Myanmar. This trial observed a protective effect against *P. falciparum* but not *P. vivax* infection (13), suggesting that repellent prevents new infections transmitted through the bites of infective *Anopheles* mosquitoes rather than relapsing *P. vivax* infections from dormant hypnozoites. Within this cohort, we measured antibodies against the established *Anopheles* salivary antigen gSG6-P1 and determined the effect of the distribution of repellent on the antibody response to assess the utility of this biomarker as a surrogate outcome for *Anopheles* biting in vector control intervention efficacy and effectiveness studies.

## Methodology

Samples and data were obtained from a stepped-wedge cluster randomised controlled trial assessing the effectiveness of personal insect repellent delivered by VHVs to 114 villages in Southeast Myanmar. Details of this study design has been published previously (13, 14). Briefly, villages were randomised into 14 blocks, which transitioned from a control to an intervention state in random sequence monthly, across a 15-month period so that all villages eventually received repellent; with rapid diagnostic tests (RDT) and dried blood spots (DBS) also collected monthly. Repellent was distributed to all villagers (residents, migrants and forest dwellers) at community meetings held by VHVs and local field implementation staff, where they received verbal explanations of how repellent works and its proper application. Villagers were asked to return empty repellent tubes to VHVs (although not a requirement) for replacement to ensure continued coverage. Informed consent was collected from all participants or parents/guardians, and ethics was provided by the Government of the Republic of the Union of Myanmar Ministry of Health Department of Medical Research and the Alfred Hospital Ethics Committee. This trial is registered in the Australian New Zealand Clinical Trials Registry (ACTRN12616001434482; approved retrospectively 14 October 2016).

### *Plasmodium* spp. infection

*P. falciparum* and *P. vivax* infection were detected by SD bioline *Pf/Pv* combo RDT performed by malaria volunteers in the field. Participants were also invited to provide a DBS for molecular detection of parasitaemia and immunoassays. DNA was extracted and tested for *P. vivax* and *P. falciparum* using a duplex qPCR as previously reported (13).

### Immunoassays

Detection of total IgG against *Anopheles* gambiae salivary gland 6 peptide gSG6-P1 by enzyme-linked immunosorbent assay (ELISA) was performed using a liquid handling robot (JANUS automated work station, Perkin Elmer) according to a high-throughput protocol described in Kearney *et al*. (15).

### Measures

#### Outcomes

The primary outcomes were the levels (continuous OD 405nm) and seropositivity (binary: defined as having an OD greater than the mean plus three standard deviations of unexposed controls from Melbourne, Australia) of total IgG antibodies against gSG6-P1. Antibodies were measured in each individual for whom a DBS was collected using the protocol outlined above.

#### Exposure and covariates

To estimate the effect of repellent on antibodies to gSG6-P1, repellent distribution was included as a binary time-varying (monotonic) variable that changed from a control to intervention state for a given cluster of villages at a specific month over the 15-month study. To model long term cumulative effects of repellent distribution on antibodies (to allow antibody decay), we generated a series of variables representing lagged effects of repellent distribution (by generating lags of monthly increments up to 7-months [half the length of the study]). While individual-level data on the frequency of repellent use was not collected, for the purpose of interpretation, we refer to these cumulative effects as a period of repellent use (*i*.*e*. a 6-month lag is interpreted as 6-months of repellent use). Additional covariates of interest included participant risk group (village resident, migrant, forest dweller), seasonality (categorical: hot, rainy, cool), study-level time (continuous: month) and participant-level time (continuous: months since first test).

### Statistical analysis

To determine the effect of distribution of personal repellent on longitudinal measurements of antibody levels (continuous) or seropositivity (binary), mixed-effects linear and logistic regression modelling was performed, respectively, adjusting for covariates of interest and including crossed random effects (*i*.*e*. intercept) for village (level-2) and study-level time (level-2) heterogeneity in antibodies to gSG6-P1 and a random effect (*i*.*e*. slope) for village-specific heterogeneity in the effect of the repellent intervention. A series of models estimating the cumulative effects of repellent distribution were also estimated to quantify possible long-term effects of repellent use on antibody decay. As we identified *a priori* that the effect of repellent on antibodies to gSG6-P1 was likely to vary between participant risk group, we included an interaction term between risk group and repellent distribution. In addition to the cross-random effects mixed modelling, we also sought to account for any individual level antibody dynamics. We estimated the effect of repellent (instantaneous and cumulative) on gSG6-P1 IgG levels for each risk group in mixed-effects linear regression with nested random effects to account for repeated measures, with samples (level-1) nested in participants (level-2) nested in villages (level-3). In addition to an interaction term between the intervention (instantaneous and cumulative) and participant risk group, the model adjusts for covariates of interest including seasonality, study- and participant-level time (*i*.*e*. month of study and months from first test).

Empirical Bayes best linear unbiased predictions (BLUPs) were used to estimate village-level random slopes and village-specific average effect of repellent for village residents, migrants and forest dwellers. All analyses were performed using STATA (version 15.1).

## Results

After one month of all villages in the control phase (baseline), clusters of 8-12 villages transitioned monthly in a stepped-wedge manner to receive the repellent intervention, with RDTs and DBS samples collected by VHVs. A total of 14,128 DBS from 10,857 consenting participants residing in 111 of the 114 villages were available for analysis (Supplementary Figure 1). Of the individuals who provided more than one sample, most provided only a second sample (65%; 1,340/2,302) but one participant provided 10 samples. The median time between samples was 3 months (interquartile range (IQR): 2-6 months). The median age of participants was 20 years (IQR: 10-35) and 49.5% (6994/14128) were male (Table1). Village residents contributed 42.2% of DBS (5957/14128), with the remaining DBS collected from high-risk participants: forest dwellers (46.7% (6600/14128) and migrants (11.1%; 1571/14128). Overall, the prevalence of *Plasmodium* spp. detection by RDT and PCR was very low (0.1%; 20/14,128 and 3.2%; 419/13,157 respectively), while the median levels (2.1 (IQR: 1.5, 2.4)) and seroprevalence (59.9% (8462/14128) of IgG antibodies against the *Anopheles* salivary protein gSG6-P1 was high (Table1). The levels and seroprevalence of anti-gSG6-P1 IgG antibodies were dynamic over the course of the study, declining from April to June 2015 coinciding with the end of the hot and beginning of the rainy season, before rising during the rainy season from July until October 2015 and then sustained at high levels for the remainder of the study (Figure 1). These trends were similar between participant risk groups.

**Table 1.** Description of participant characteristics and malaria transmission and vector exposure outcomes.

|  | **Population with DBS**<br>**n=14,128** |
| --- | --- |
| **Number of participants, (n)** | 10,857 |
| **Subset with repeat sampling** |  |
| Number of participants, (n) | 2,032 |
| Number of samples per participant, Median (IQR) [Range] | 2 (2, 4) [2, 10] |
| Time from first sample (months), Median (IQR) [Range] | 2.2 (0, 6.4) [0, 14.5] |
| **Sex, % (n)** |  |
| Male | 49.5 (6,994) |
| Female | 50.5 (7,134) |
| **Age (years), Median (IQR) [Range]** | 20.0 (10.0, 35.0) [0.1, 91.0] |
| **Risk group, % (n)** |  |
| Village Resident | 42.2 (5,957) |
| Migrant | 11.1 (1,571) |
| Forest Dweller | 46.7 (6,600) |
| **State, % (n)** |  |
| Bago (East) | 11.2 (1,581) |
| Kayah | 27.4 (3,872) |
| Kayin | 61.4 (8,675) |
| **RDT, % (n)** |  |
| Negative | 99.9 (14108) |
| *Pf*<sup>+</sup> | <0.1 (7) |
| Non-*Pf*<sup>+</sup> | <0.1 (12) |
| Mixed | <0.1 (1) |
| **PCR, % (n)** | **n=13157** |
| Negative | 96.8 (12,738) |
| *Pf*<sup>+</sup> | 1.6 (207) |
| *Pv*<sup>+</sup> | 0.9 (123) |
| Mixed | 0.7 (89) |
| **gSG6-P1 IgG** |  |
| Seropositivity, % (n) | 59.9 (8,462) |
| Levels, Median (IQR) [Range] | 2.1 (1.5, 2.4) [0, 3.1] |
| Levels, Mean (SD) | 1.9 (6.5) |

IQR – inter-quartile range (given as the 25<sup>th</sup> and 75<sup>th</sup> percentiles); RDT – rapid diagnostic test; PCR – polymerase chain reaction; SD – standard deviation

**Figure 1.**
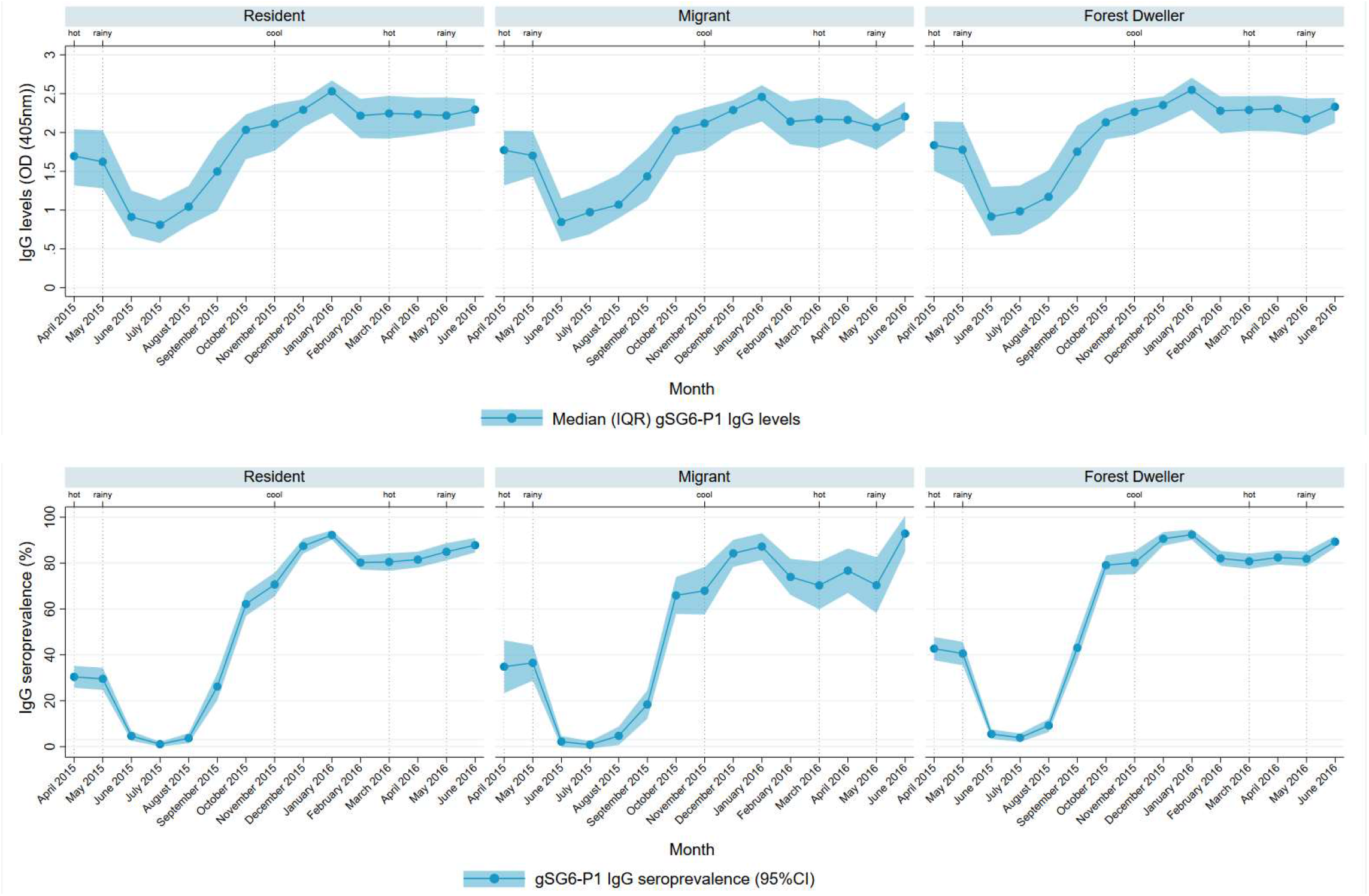
Longitudinal dynamics of anti-gSG6-P1 IgG antibodies for each participant risk group. Figure shows the median (inter-quartile range (IQR)) of the levels and the seroprevalence (95% confidence interval (95%CI)) of IgG antibodies against gSG6-P1 for village residents, migrants and forest dwellers.

### The cumulative effects of long-term repellent use suggest a protective effect on gSG6-P1 IgG antibody levels, especially for high-risk participants

The overall instantaneous effect of village-level distribution of personal insect repellent on anti-gSG6-P1 IgG antibody levels and seropositivity was: a mean difference of 0.005 (95%CI: −0.03, 0.04; *p*=0.784) and a relative change in odds (i.e. Odds Ratio) of 1.07 (95%CI: 0.88, 1.30; *p*=0.490), respectively (Figure 2). Participant risk group moderated the effect of repellent use on gSG6-P1 IgG antibodies levels but not seropositivity (*p*-value for interaction term between risk group and the instantaneous effect of the intervention: *p*=0.001 and *p*=0.110, respectively; Supplementary Table **2**), corresponding to an increase of 0.04 units (95%CI: 0.00, 0.08; *p*=0.042) in anti-gSG6-P1 IgG levels associated with the instantaneous effect of repellent for village residents, and respective decreases of 0.03 (95%CI: −0.09, 0.02; *p*=0.272) and 0.02 units (95%CI: −0.05, 0.02; *p*=0.404) in anti-gSG6-P1 IgG levels for migrants and forest dwellers (Figure 3).

**Figure 2.**
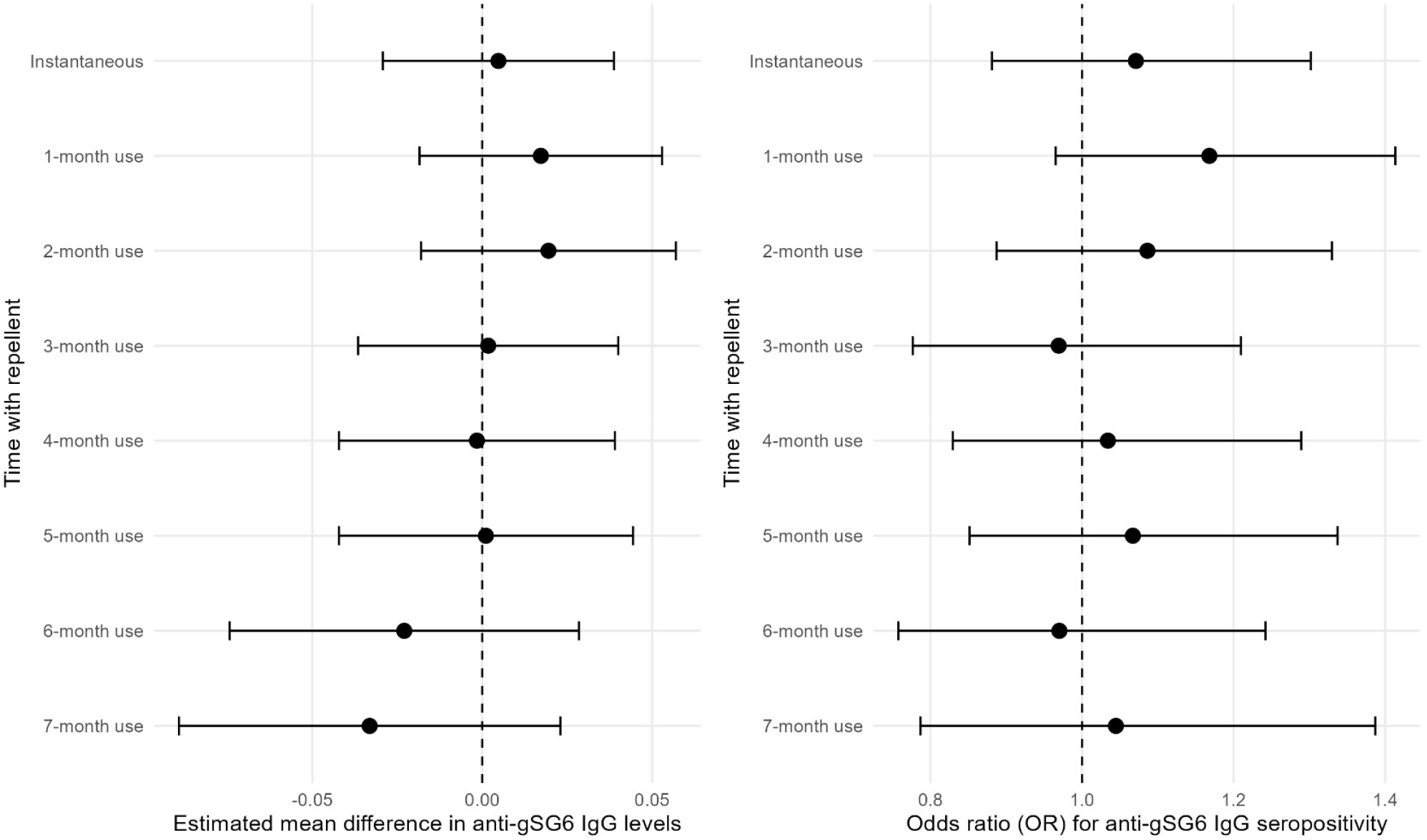
Forest plots showing the instantaneous and cumulative effects of repellent distribution on anti-gSG6-P1 IgG antibody levels and odds of seropositivity in all participants. Figures show the estimated mean difference (95% confidence interval (95%CI)) in anti-gSG6-P1 IgG antibody levels and estimated odds ratios (95%CI) of anti-gSG6-P1 IgG seropositivity for distributed repellent compared to the period of no exposure to repellent. Estimates were generated using mixed effects linear and logistic modelling respectively, adjusting for temporality, season and risk group, with crossed random-effects for village, month and village-specific heterogeneity in the effect of repellent. Separate models were fitted to estimate the instantaneous and cumulative effects of repellent distribution (1-7 months). n=14,128 antibody measurements from 10,857 participants.

**Figure 3.**
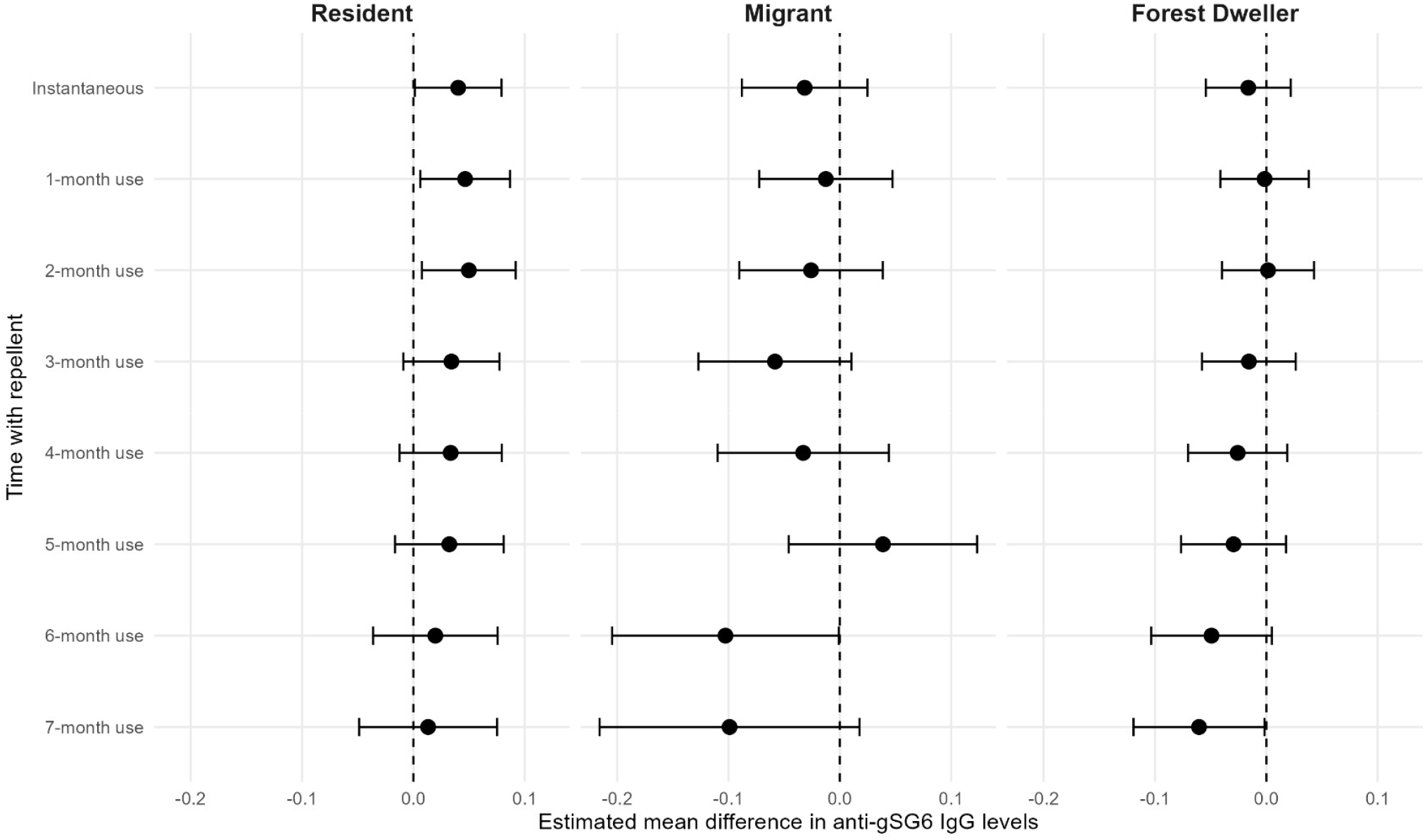
Forest plots showing the instantaneous and cumulative effects of repellent distribution on anti-gSG6-P1 IgG antibody levels for each risk group. Figures show the estimated mean difference (95% confidence interval) in anti-gSG6-P1 IgG antibody levels for distributed repellent compared to the period of no exposure to repellent separately for village residents, migrants and forest dwellers. Estimates were generated using mixed effects linear modelling adjusting for temporality, season and the moderating effect of risk group, with crossed random-effects for village, month and village-specific heterogeneity in the effect of repellent. Separate models were fitted to estimate the instantaneous and cumulative effects of repellent distribution (1-7 months). n=14,128 antibody measurements from 10,857 participants.

While no instantaneous effect of repellent on anti-gSG6-P1 IgG antibody levels was observed, when considering cumulative effects from longer term use there was a trend towards reduced anti-gSG6-P1 IgG levels overall (Figure 2) and for high-risk groups of migrants and forest dwellers (Figure 3). For example, 6-months of repellent distribution was associated with a small decrease (−0.02; 95%CI: −0.07, 0.03; *p=*0.381) in anti-gSG6-P1 IgG levels overall (Figure 2). However, for migrants and forest dwellers there were decreases of 0.10-units (95%CI: −0.20, −0.001; *p*=0.048) and 0.05-units (95%CI: −0.10, 0.005; *p*=0.075) in anti-gSG6-P1 IgG levels associated with 6-months of repellent distribution respectively, compared to a small increase of 0.02-units (95%CI: −0.04, 0.08; *p*=0.489) in anti-gSG6-P1 IgG levels for village residents (Figure 3). However, the confidence intervals for the instantaneous and cumulative effects 1-to 5-months of distribution are wide, indicating that the evidence for these associations are weak. A similar although less consistent pattern of reduced antibody levels was observed in high-risk groups when anti-gSG6-P1 IgG seropositivity was considered as the outcome (Supplementary Figure 2). Of note, the inclusion of a random effect for participant to account for variation in antibody levels at the individual level did not explain any additional variability in anti-gSG6-P1 IgG antibody level (Supplementary Figure 3).

### Significant heterogeneity in the effect of repellent on gSG6-P1 IgG between villages

We observed significant heterogeneity in the effect of repellent (instantaneous and cumulative) on anti-gSG6-P1 IgG levels between villages (Supplementary Figure 4). For example, the 95% reference range of village-specific effects for 6-months of repellent distribution ranged from a 0.24-unit decrease to a 0.28-unit increase in anti-gSG6-P1 IgG levels for village residents, a 0.36-unit decrease to a 0.16-unit increase for forest dwellers and a 0.31-unit decrease to a 0.21-unit increase for migrants (Supplementary Figure 5).

## Discussion

Detection of antibodies specific for *Anopheles* salivary antigens has the potential to be an innovative approach to measure vector exposure and may have application as a surrogate outcome in vector control intervention effectiveness trials. This study demonstrates that the effect of repellent distribution on decreasing anti-gSG6-P1 IgG antibodies differed according to a participant’s risk group and we observed large variation in the village-specific effects of repellent on anti-*Anopheles* saliva antibody levels. Importantly, our study is the first to quantify the instantaneous and cumulative effects of an intervention on antibodies to *Anopheles* salivary proteins, providing valuable insights on antibody longevity and village level heterogeneity. While our study highlights the potential utility of these antibodies as a serosurveillance tool, it also provides important parameters to inform the potential use and study design for salivary antibodies as candidate surrogate endpoints in future vector control intervention effectiveness trials.

This study identified that the effect of repellent distribution on *Anopheles* salivary antibodies was variable and changed over time. While no instantaneous association between the distribution of repellent and anti-gSG6-P1 antibodies was observed, consideration of a series of lags to model the cumulative effect of repellent use saw some evidence of a reduction in antibody levels at 6- and 7-months after distribution. As antibodies are dynamic and wane in the absence of ongoing antigenic exposure (*i*.*e*. mosquito biting), the estimation of a series of lags allows for quantification of gradual antibody decay induced by sustained reductions in mosquito bites from cumulative repellent use. As the first study to employ this approach, we provide important parameters on antibody longevity and suggest that intervention effectiveness trials such as these would need to follow participants for at-least 6 months to adequately capture antibody decay. These estimates are in line with the estimated the anti-SG6 IgG half-life of ~200-400 days following biting (depending on the species-specific antigen) (16). Of note, we only investigated lags of up to the half-way point of the study (7-months) as the stepped-wedge trial design saw fewer samples available in the intervention state in latter periods of the trial (*i*.*e*. only 1,555 samples available from 1,399 individuals with 8-months of repellent distribution: 69 from migrants, 908 for forest dwellers and 578 from residents), which may cause us to estimate the effect of the intervention with reduced precision. Importantly this study identified that the reduction in antibody levels associated with the intervention was observed for migrants and forest dwellers, but not for village residents. This greater protective effect for the high-risk groups may be due to greater overall exposure to *Anopheles* bites (and therefore a greater magnitude of protection from exposure) compared to village residents, or perhaps higher rates of repellent use amongst these at-risk groups. Of note, while repellent was rolled out to all study participants regardless of risk-group status, participants were specifically encouraged to use repellent when travelling into forested areas. Yet as neither entomological endpoints nor individual-level data on the frequency of repellent use were collected in this study, we are unable to definitively quantify the associations between serological endpoints and observed mosquito biting nor be certain of the drivers of these risk-group differences. Regardless, our findings of a trend of reduced *Anopheles* salivary antibody levels for migrants and forest dwellers after allowing for antibody decay (although small in magnitude) suggest that personal repellent may serve as an effective supplementary intervention in preventing mosquito biting and residual transmission in these high-risk groups.

Overall, the magnitude of the protective effect of repellent on antibody levels was relatively small with wide confidence intervals and varied significantly between villages, with repellent distribution associated with reductions in anti-gSG6-P1 IgG levels in some villages but increases in others. This has implications for the reliability and external validity of this approach to assess the effectiveness of an intervention. As this weak evidence of an effect could be due an overall weak or quickly reversed effect of repellent on *Anopheles* exposure in our study (although not observed in the *P. falciparum* results (13)), perhaps different vector control interventions with different modes of action and often greater magnitudes of protective effects (e.g. long-lasting insecticide treated nets, indoor residual spraying, spatial emanators or outdoor residual insecticide spraying(17, 18)) may correlate with greater reductions in antibody levels. Our study also identified that there was no association between repellent distribution and anti-gSG6-P1 antibodies when seropositivity was considered as an outcome. This is potentially due to a reduction in statistical power through the dichotomisation of the continuous outcome (antibody levels), suggesting that a quantitative tool that measures antibodies as a continuous outcome would likely be more informative as an outcome in vector control intervention effectiveness trials. Furthermore, this study identified no additional benefit of quantifying the effect of the intervention on anti-gSG6-P1 antibodies at an individual-, rather than population-, level. This is in line with findings from Bottomley *et al*. (19) who indicate that clustering at the higher level is often sufficient. As most participants in our study contributed only one sample to the analysis, and individual data on repellent use was not directly observed, inferences of biting exposure and repellent efficacy may be restricted to the population level. Studies with rich longitudinal follow-up from a single study site that assess the effect of the intervention at the individual level (20, 21) may be better placed to assess individual level changes over time.

Key strengths of the present study relate to the size of the sample, the duration of follow-up and the stepped-wedge trial design. Having repeat sampling of each village in both the control and intervention states has allowed for estimation of not only the instantaneous, but also the cumulative effects of repellent distribution on antibody levels for the first time. Consideration of antibody levels and seroprevalence as a trial outcome has greatly increased the number of events (n=14,128 for antibody levels and n=8,462 for seropositive samples, compared to RDT (n=20) or PCR (n=419) detected *Plasmodium* spp. infection), which may permit estimation of repellent effectiveness with increased precision. This is highly relevant for settings with low transmission and approaching elimination, when detection of the few positive cases becomes increasingly challenging. However, the ability of these *Anopheles* salivary antibodies to serve as surrogate trial outcomes are dependent on how well they correlate with malarial and entomological outcomes. This is typically assessed in a meta-analytical framework involving a set of studies investigating the same intervention that collects both the surrogate and true endpoints (22). Such an investigation could be the subject of future work, however, most trials that have investigated these antibody biomarkers have been largely descriptive and highly heterogeneous across population, trial design, antibody measurement and primary endpoint measure.

A limitation of the study is that individual repellent use was not directly observed. This precludes the assessment of the use of antibodies specific for *Anopheles* salivary antigens to assess intervention compliance and limits the ability to infer whether differences in antibody levels between risk groups was due to better compliance by migrants and forest dwellers. Individually randomised designs that adequately observe adherence may be better placed to determine the levels of personal (rather than community) protection, however, these study designs are challenging to perform at scale. Furthermore, because *Anopheles*-biting exposure data was not measured at either the individual- or population-level, the rate of antibody boosting and decay in response to the presence and absence of biting exposure could not be estimated. Future studies could also explore the application of additional salivary antibody biomarkers with different dynamic profiles (e.g. species-specific antigens or antibody isotypes/subclasses with shorter half-lives) to determine the best serological biomarker of recent changes in *Anopheles* biting exposure.

## Conclusions

These findings provide evidence that human antibodies to *Anopheles* salivary proteins have potential as a surrogate outcome in vector control intervention effectiveness trials in the GMS as an alternative to impractical and insensitive human landing catches and in areas with low disease rates. We provide the first estimates of the delayed effects of an intervention on anti-gSG6-P1 IgG antibody levels to allow modelling of gradual antibody decay, suggesting that future vector control intervention effectiveness trials would likely need to be longitudinal in design, following participants for at-least 6-months. However, further studies that measure the effects of other vector control interventions and quantify changes in these antibodies over time are needed to inform if these anti-*Anopheles* salivary antibodies can be appropriately applied as a surrogate outcome measure in vector control intervention effectiveness trials.

## Acknowledgements

We would like to thank the local communities and village health volunteers for their participation in the research; Karuna Mission Social Solidarity (KMSS) staff Win Tun Kyi, Augustine Tual Sian Piang, Min Thant Zin Latt, Nwe Ni Aye, and Khine Zar Lwin (KMSS National Office, Yangon, Myanmar); Joseph Maung Win, Tun Tun Aung, Richard Joseph, Shein Thu Ag, Thet Naing, and Myo Tint (KMSS Yangon, Yangon, Myanmar); Ludovico Saw Piko, Benedetta, Win Win Aye, Ngo Petru, Maurice Nyo, Daniel Win, and Saw Golbert (KMSS Taunggo, Taunggo, Myanmar); Albino Htwe Win, Tin Aung, Perpetua Aye Aye Mon, Mupaula, John Bosco, Alfred, John Min Aung, and Poe Rah (KMSS Loikaw, Loikaw, Myanmar); Paul Thar San, Saw Isidore, Theresa, Saw Micheal, and Saw Pho Muo (KMSS Hpa An, Hpa An, Myanmar) for the local advocacy, coordination, and field work; Burnet Institute staff Toe Than Tun, Phone Myint Win, Poe Poe Aung, Ai Pao Yawn, Kaung Myat Thu, May Chan Oo, Galau Naw Hkaung, Ei Phyu Htwe, Aung Khine Zaw, Lia Burns, Naanki Pasricha, Ricardo Ataíde, James Beeson, Brendan Crabbe and Nicole Romero for contributing toward technical, coordination, and management support and contextual inputs; Alissa Robertson and Josh Charles for the assistance in sample processing; Victor Chaumeau for support and entomological expertise; and Myanmar Ministry of Health staff Late Dr. Thandar Lwin, Dr. Thaung Hlaing, Dr. Than Naing Soe, and Dr. Kyawt Mon Win for the technical and administrative support.

## Data, Materials, and Software Availability

Data cannot be made publicly available because it would breach compliance with the ethical framework of the Ethics Review Committee on Medical Research Involving Human Subjects, Department of Medical Research, Myanmar Ministry of Health and Sports. Deidentified individual participant data are stored on Burnet Institute servers and will be made available from the corresponding author to applicants who provide a sound proposal to The Ethics Review Committee on Medical Research Involving Human Subjects, Department of Medical Research, Myanmar Ministry of Health and Sports (No. 5 Ziwaka Road, Dagon PO Yangon, Myanmar; (+95) 01 375447 extension 118) contingent of their approval.

