## Supplementary tables and figures for "*Anopheles* salivary antibody biomarkers as surrogate outcomes measures to assess the effectiveness of topical repellent in Southeast Myanmar"

[1] Centre for Epidemiology and Biostatistics, Melbourne School of Population and Global Health, The University of Melbourne, Melbourne, VIC, Australia, [2] Disease Elimination Program, Burnet Institute, Melbourne, VIC, Australia, [3] Biostatistics Unit, Faculty of Health, Deakin University, Melbourne, Australia, [4] Health Security and Malaria Program, Burnet Institute Myanmar, Yangon, Myanmar, [5] Department of Medicine at the Doherty Institute, The University of Melbourne, Melbourne, VIC, Australia, [6] Institute for Physical and Mental Health and Clinical Translation (IMPACT), School of Medicine, Deakin University, Geelong, VIC, Australia, [7] Department of Public Health, Myanmar Ministry of Health and Sports, Nay Pyi Taw, Myanmar, [8] Centre for Tropical Medicine and Global Health, Nuffield Department of Medicine, University of Oxford, Oxford, UK, [9] Department of Epidemiology and Preventive Medicine, Monash University, Melbourne, VIC, Australia

**Keywords:** salivary biomarkers, vector control trials, malaria, Anopheles, surrogate outcomes, repellent.

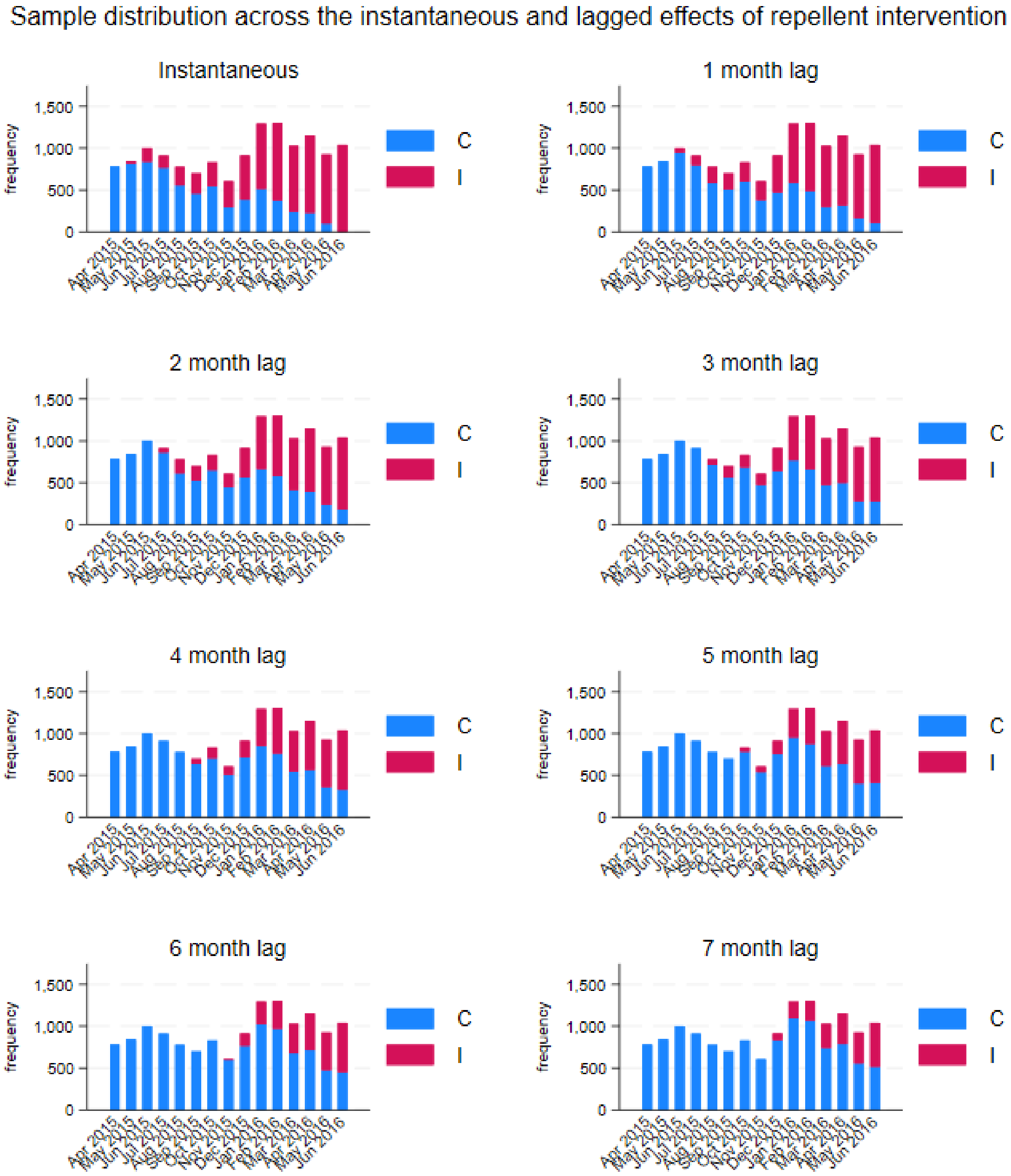


**Supplementary Figure 1. Distribution of samples collected over time, coloured by intervention state for the instantaneous and lagged effects of repellent distribution.** Shows the frequency of samples collected per month, with blue representing the control period (C), and red indicating the intervention (I; *i.e.,* repellent) period.

**Supplementary Table 1. Instantaneous association between village-level distribution of repellent and anti-gSG6-P1 IgG levels and seropositivity (n=14,128 antibody measurements from N=10,857 participants).**

| Variable | Mean difference in anti-gSG6-P1 IgG level | | | | Odds ratio for anti-gSG6-P1 IgG seropositivity | | | |
|---|---|---|---|---|---|---|---|---|
| | **MD** | **95%CI** | | ***p*** | **OR** | **95%CI** | | ***p*** |
| *Intervention* | | | | | | | | |
| No repellent | Ref. | | | | Ref. | | | |
| Repellent | 0.00 | -0.03 | 0.04 | 0.784 | 1.07 | 0.88 | 1.30 | 0.490 |
| *Risk group* | | | | **<0.001* | | | | **0.001* |
| Village Resident | Ref. | | | | Ref. | | | |
| Migrant | -0.02 | -0.05 | 0.01 | 0.153 | 0.85 | 0.72 | 1.01 | 0.063 |
| Forest Dweller | 0.05 | 0.03 | 0.07 | <0.001 | 1.21 | 1.08 | 1.36 | 0.001 |
| *Month* | 0.06 | 0.03 | 0.09 | <0.001 | 1.35 | 1.18 | 1.54 | 0.000 |
| *Season* | | | | **0.005* | | | | **0.014* |
| Cool | Ref. | | | | Ref. | | | |
| Hot | -0.14 | -0.51 | 0.22 | 0.437 | 0.45 | 0.09 | 2.30 | 0.338 |
| Rainy | -0.47 | -0.77 | -0.17 | 0.002 | 0.14 | 0.04 | 0.54 | 0.004 |
| **Random effects variances** | **var** | | | | **var** | | | |
| *Intercept: Month* | 0.06 | | | | 1.17 | | | |
| *Intercept: Village* | 0.02 | | | | 0.38 | | | |
| *Slope: Intervention* | 0.01 | | | | 0.26 | | | |

Table shows estimated mean difference (MD) and odds ratio (OR), 95% confidence interval (95%CI), p-value (*p*), with random-effects variances (var).

[a] Mixed effects linear and logistic modelling estimating the association between the distribution of repellent and anti-gSG6-P1 IgG levels and seropositivity respectively, adjusting for temporality, season and risk group, with random-effects for village, month and village-specific heterogeneity in the effect of repellent.

*Indicates p-value for Wald joint tests for variables with >2 categories (risk group and season)

**Supplementary Table 2. Instantaneous association between village-level distribution of repellent and anti-gSG6-P1 IgG levels and seropositivity, moderated by risk group (n=14,128 antibody measurements from N=10,857 participants).**

| | **Mean difference in anti-gSG6-P1 IgG level** | | | | **Odds ratio for anti-gSG6-P1 IgG seropositivity** | | | |
|---|---|---|---|---|---|---|---|---|
| **Variable** | **MD** | **95%CI** | | ***p*** | **OR** | **95%CI** | | ***p*** |
| *Intervention* | | | | | | | | |
| No repellent | Ref. | | | | Ref. | | | |
| Repellent | 0.04 | 0.00 | 0.08 | 0.042 | 1.19 | 0.95 | 1.50 | 0.138 |
| *Risk group* | | | | **<0.001* | | | | **<0.001* |
| Village Resident | Ref. | | | | Ref. | | | |
| Migrant | 0.01 | -0.02 | 0.05 | 0.552 | 0.86 | 0.69 | 1.08 | 0.186 |
| Forest Dweller | 0.08 | 0.05 | 0.10 | 0.000 | 1.36 | 1.16 | 1.59 | <0.001 |
| *Intervention by risk group* | | | | **0.001* | | | | **0.110* |
| Village Resident | Ref. | | | | Ref. | | | |
| Migrant | -0.07 | -0.13 | -0.02 | 0.009 | 0.99 | 0.71 | 1.38 | 0.944 |
| Forest Dweller | -0.06 | -0.09 | -0.02 | 0.001 | 0.79 | 0.64 | 0.99 | 0.043 |
| *Month* | 0.06 | 0.03 | 0.09 | 0.000 | 1.35 | 1.18 | 1.54 | <0.001 |
| *Season* | | | | **0.005* | | | | **0.014* |
| Cool | Ref. | | | | Ref. | | | |
| Hot | -0.14 | -0.51 | 0.22 | 0.437 | 0.45 | 0.09 | 2.31 | 0.340 |
| Rainy | -0.47 | -0.77 | -0.17 | 0.002 | 0.14 | 0.04 | 0.55 | 0.005 |
| **Random effects variances** | **var** | | | | **var** | | | |
| *Intercept: Month* | 0.06 | | | | 1.18 | | | |
| *Intercept: Village* | 0.02 | | | | 0.38 | | | |
| *Slope: Intervention* | 0.01 | | | | 0.26 | | | |

Table shows estimated mean difference (MD) and odds ratio (OR), 95% confidence interval (95%CI), p-value (*p*), with random-effects variances (var).

[a] Mixed effects linear and logistic modelling estimating the association between the distribution of repellent and anti-gSG6-P1 IgG levels and seropositivity respectively, adjusting for temporality, season and the moderating effect of risk group, with random-effects for village, month and village-specific heterogeneity in the effect of repellent.

*Indicates p-value for Wald joint tests for variables with >2 categories (risk group and season)

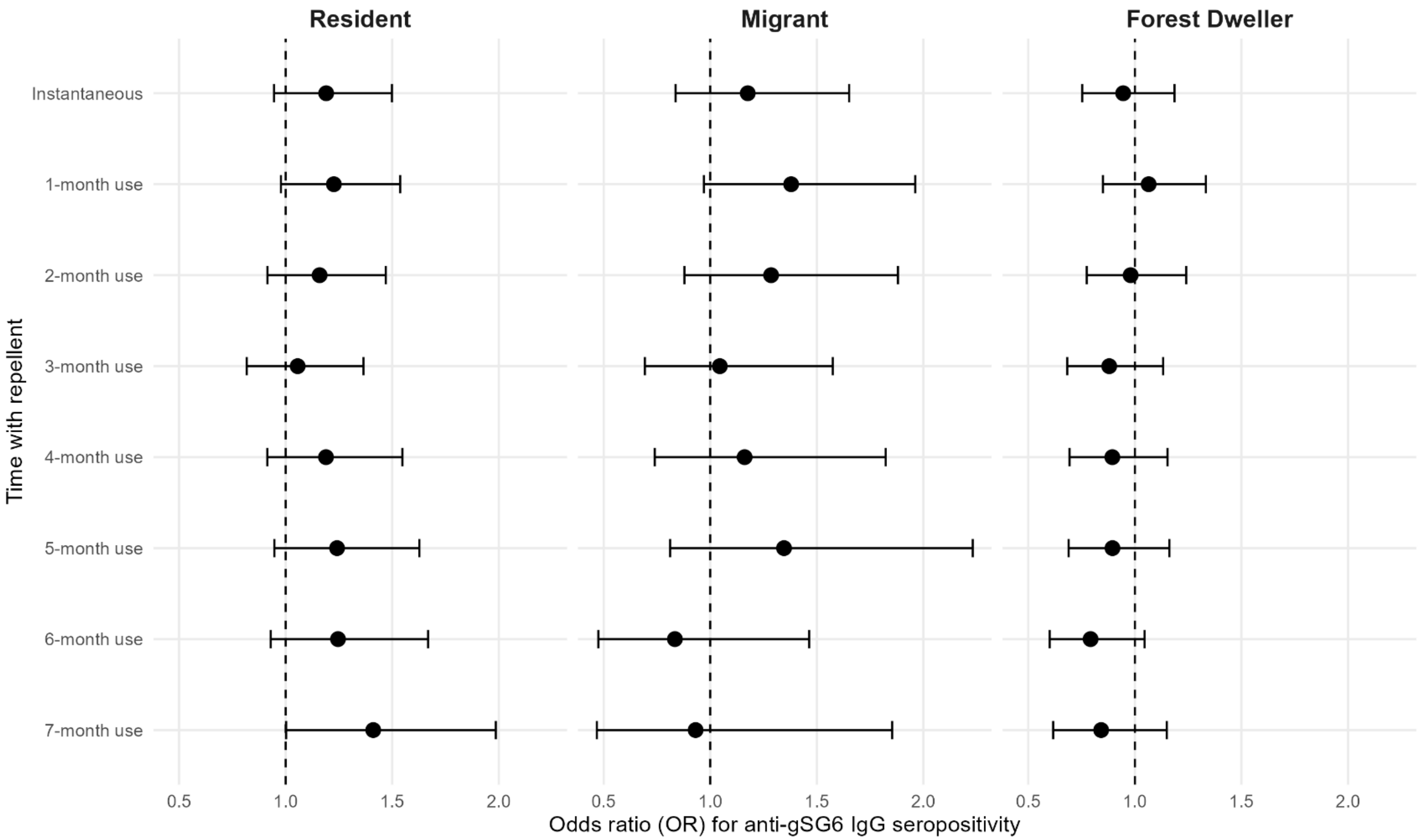


**Supplementary Figure 2. Forest plots showing the instantaneous and cumulative effects of repellent use on the odds of anti-gSG6-P1 IgG seropositivity for each risk group.** Figures show the estimated odds ratios (95% confidence interval) of anti-gSG6-P1 IgG seropositivity for repellent use compared to the period of no exposure to repellent separately for village residents, migrants and forest dwellers. Estimates were generated using mixed effects logistic modelling adjusting for temporality, season and the moderating effect of risk group, with crossed random-effects for village, month and village-specific heterogeneity in the effect of repellent. Separate models were fitted to estimate the instantaneous and cumulative effects of repellent use (1-7 months of use). n=14,128 antibody measurements from N=10,857 participants.

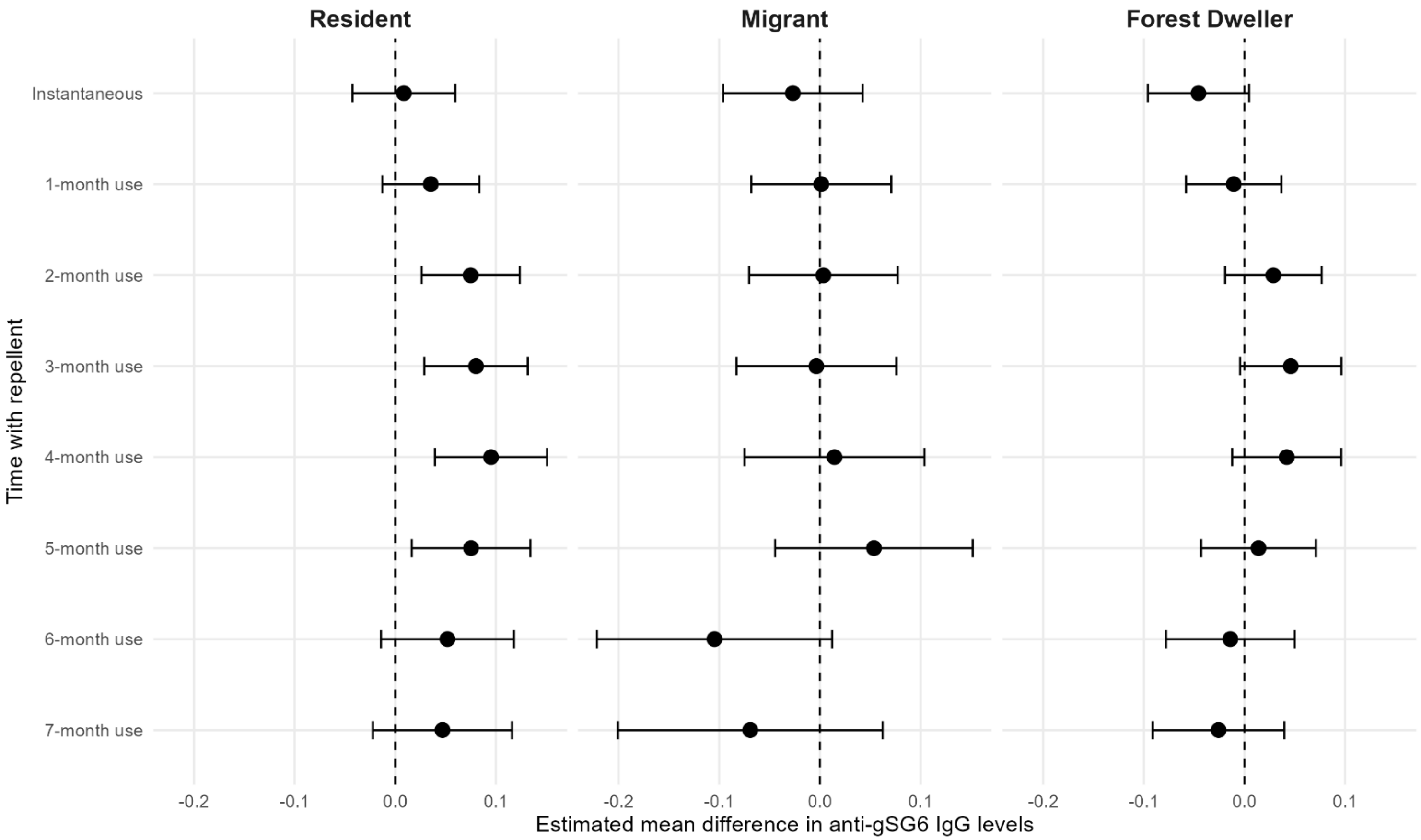


**Supplementary Figure 3. Forest plots showing the instantaneous and cumulative effects of repellent use on anti-gSG6-P1 IgG antibody levels for each risk group, after adjusting for individual level antibody dynamics.** Figures show the estimated mean difference (95% confidence interval) in anti-gSG6-P1 IgG antibody levels for repellent use compared to the period of no exposure to repellent separately for village residents, migrants and forest dwellers. Estimates were generated using mixed effects linear modelling adjusting for study- and individual-level temporality, season and the moderating effect of risk group, with nested random-effects to account for repeated measures (with samples nested in individuals nested in villages) and village-specific heterogeneity in the effect of repellent. Separate models were fitted to estimate the instantaneous and cumulative effects of repellent use (1-7 months of use). n=14,128 antibody measurements from N=10,857 participants.

**Supplementary Table 3. Cumulative effects of repellent use on gSG6-P1 IgG antibody levels (n=14,128 antibody measurements from N=10,857 participants).**

| Variable | 1 month use | | | | 2 months use | | | | 3 months use | | | | 4 months use | | | |
|---|---|---|---|---|---|---|---|---|---|---|---|---|---|---|---|---|
| | **MD** | **95%CI** | | ***p*** | **MD** | **95%CI** | | ***p*** | **MD** | **95%CI** | | ***p*** | **MD** | **95%CI** | | ***p*** |
| *Intervention* | | | | | | | | | | | | | | | | |
| No repellent | Ref. | | | | Ref. | | | | Ref. | | | | Ref. | | | |
| Repellent | 0.02 | -0.02 | 0.05 | 0.344 | 0.02 | -0.02 | 0.06 | 0.308 | 0.00 | -0.04 | 0.04 | 0.928 | 0.00 | -0.04 | 0.04 | 0.940 |
| *Risk group* | | | | *<0.001 | | | | *<0.001 | | | | *<0.001 | | | | *<0.001 |
| Village Resident | Ref. | | | | Ref. | | | | Ref. | | | | Ref. | | | |
| Migrant | -0.02 | -0.04 | 0.01 | 0.231 | -0.02 | -0.05 | 0.01 | 0.166 | -0.02 | -0.05 | 0.01 | 0.146 | -0.02 | -0.05 | 0.01 | 0.175 |
| Forest Dweller | 0.05 | 0.03 | 0.07 | <0.001 | 0.05 | 0.03 | 0.07 | <0.001 | 0.05 | 0.04 | 0.07 | <0.001 | 0.05 | 0.03 | 0.07 | <0.001 |
| *Month* | 0.06 | 0.03 | 0.09 | <0.001 | 0.06 | 0.03 | 0.09 | <0.001 | 0.06 | 0.03 | 0.09 | <0.001 | 0.06 | 0.03 | 0.09 | <0.001 |
| *Season* | | | | *0.006 | | | | *0.005 | | | | *0.005 | | | | *0.005 |
| Cool | Ref. | | | | Ref. | | | | Ref. | | | | Ref. | | | |
| Hot | -0.15 | -0.51 | 0.22 | 0.436 | -0.14 | -0.51 | 0.22 | 0.437 | -0.14 | -0.51 | 0.22 | 0.440 | -0.15 | -0.51 | 0.22 | 0.434 |
| Rainy | -0.47 | -0.77 | -0.17 | 0.002 | -0.47 | -0.77 | -0.17 | 0.002 | -0.47 | -0.77 | -0.17 | 0.002 | -0.47 | -0.77 | -0.17 | 0.002 |
| **Random effect variances** | | | | | | | | | | | | | | | | |
| *Intercept: Month* | | | | 0.06 | | | | 0.06 | | | | 0.06 | | | | 0.06 |
| *Intercept: Village* | | | | 0.02 | | | | 0.02 | | | | 0.02 | | | | 0.02 |
| *Slope: Intervention* | | | | 0.01 | | | | 0.02 | | | | 0.02 | | | | 0.02 |

Table shows estimated mean difference (MD), 95% confidence interval (95%CI), p-value (p), with random-effects variances. Mixed effects linear modelling estimating the association between the distribution of repellent and anti-gSG6-P1 IgG levels, adjusting for temporality, season and risk group, with random-effects for village, month and village-specific heterogeneity in the effect of repellent.

*Indicates p-value for Wald joint tests for variables with >2 categories (risk group and season)

| Variable | 5 months use | | | | 6 months use | | | | 7 months use | | | |
|---|---|---|---|---|---|---|---|---|---|---|---|---|
| | **MD** | **95%CI** | | ***p*** | **MD** | **95%CI** | | ***p*** | **MD** | **95%CI** | | ***p*** |
| **Fixed part** | | | | | | | | | | | | |
| *Intervention* | | | | | | | | | | | | |
| No repellent | Ref. | | | | Ref. | | | | Ref. | | | |
| Repellent | 0.00 | -0.04 | 0.04 | 0.961 | -0.02 | -0.07 | 0.03 | 0.381 | -0.03 | -0.09 | 0.02 | 0.247 |
| *Risk group* | | | | *<0.001 | | | | *<0.001 | | | | *<0.001 |
| Village Resident | Ref. | | | | Ref. | | | | Ref. | | | |
| Migrant | -0.02 | -0.05 | 0.01 | 0.188 | -0.02 | -0.05 | 0.01 | 0.148 | -0.02 | -0.05 | 0.00 | 0.084 |
| Forest Dweller | 0.05 | 0.03 | 0.07 | <0.001 | 0.05 | 0.03 | 0.07 | <0.001 | 0.05 | 0.03 | 0.07 | <0.001 |
| *Month* | 0.06 | 0.03 | 0.09 | <0.001 | 0.07 | 0.04 | 0.09 | <0.001 | 0.07 | 0.04 | 0.09 | <0.001 |
| *Season* | | | | *0.005 | | | | *0.005 | | | | *0.005 |
| Cool | Ref. | | | | Ref. | | | | Ref. | | | |
| Hot | -0.15 | -0.51 | 0.22 | 0.425 | -0.15 | -0.51 | 0.22 | 0.432 | -0.14 | -0.51 | 0.22 | 0.439 |
| Rainy | -0.48 | -0.78 | -0.18 | 0.002 | -0.47 | -0.77 | -0.18 | 0.002 | -0.47 | -0.77 | -0.17 | 0.002 |
| **Random effect variance** | **var** | | | | **var** | | | | **var** | | | |
| *Intercept: Month* | 0.06 | | | | 0.06 | | | | 0.06 | | | |
| *Intercept: Village* | 0.02 | | | | 0.02 | | | | 0.02 | | | |
| *Slope: Intervention* | 0.02 | | | | 0.03 | | | | 0.03 | | | |

Table shows estimated mean difference (MD), 95% confidence interval (95%CI), p-value (*p*), with random-effects variances (var). Mixed effects linear modelling estimating the association between the distribution of repellent and anti-gSG6-P1 IgG levels, adjusting for temporality, season and risk group, with random-effects for village, month and village-specific heterogeneity in the effect of repellent.

*Indicates p-value for Wald joint tests for variables with >2 categories (risk group and season)

**Supplementary Table 4. Cumulative effects of repellent use on gSG6-P1 IgG antibody seropositivity (n=14,128 antibody measurements from N=10,857 participants).**

| Variable | 1 month use | | | | 2 months use | | | | 3 months use | | | | 4 months use | | | |
|---|---|---|---|---|---|---|---|---|---|---|---|---|---|---|---|---|
| | **OR** | **95%CI** | | ***p*** | **OR** | **95%CI** | | ***p*** | **OR** | **95%CI** | | ***p*** | **OR** | **95%CI** | | ***p*** |
| *Intervention* | | | | | | | | | | | | | | | | |
| No repellent | Ref. | | | | Ref. | | | | Ref. | | | | Ref. | | | |
| Repellent | 1.17 | 0.97 | 1.41 | 0.11 | 1.09 | 0.89 | 1.33 | 0.423 | 0.97 | 0.78 | 1.21 | 0.782 | 1.03 | 0.83 | 1.29 | 0.765 |
| *Risk group* | | | | *0.001 | | | | *0.001 | | | | *<0.001 | | | | *0.001 |
| Village Resident | Ref. | | | | Ref. | | | | Ref. | | | | Ref. | | | |
| Migrant | 0.86 | 0.73 | 1.02 | 0.091 | 0.85 | 0.72 | 1.01 | 0.064 | 0.84 | 0.71 | 1 | 0.048 | 0.84 | 0.71 | 0.99 | 0.042 |
| Forest Dweller | 1.22 | 1.08 | 1.37 | 0.001 | 1.22 | 1.08 | 1.36 | 0.001 | 1.22 | 1.09 | 1.37 | 0.001 | 1.2 | 1.07 | 1.34 | 0.002 |
| *Month* | 1.34 | 1.18 | 1.53 | 0 | 1.35 | 1.18 | 1.54 | 0 | 1.36 | 1.2 | 1.55 | 0 | 1.35 | 1.19 | 1.54 | 0 |
| *Season* | | | | *0.014 | | | | *0.014 | | | | *0.016 | | | | *0.015 |
| Cool | Ref. | | | | Ref. | | | | Ref. | | | | Ref. | | | |
| Hot | 0.46 | 0.09 | 2.32 | 0.345 | 0.46 | 0.09 | 2.32 | 0.348 | 0.48 | 0.1 | 2.39 | 0.369 | 0.46 | 0.09 | 2.32 | 0.35 |
| Rainy | 0.14 | 0.04 | 0.55 | 0.005 | 0.14 | 0.04 | 0.55 | 0.004 | 0.15 | 0.04 | 0.57 | 0.005 | 0.15 | 0.04 | 0.56 | 0.005 |
| **Random effect variances** | **var** | | | | **var** | | | | **var** | | | | **var** | | | |
| *Intercept: Month* | 1.17 | | | | 1.15 | | | | 1.15 | | | | 1.14 | | | |
| *Intercept: Village* | 0.38 | | | | 0.37 | | | | 0.35 | | | | 0.37 | | | |
| *Slope: Intervention* | 0.22 | | | | 0.29 | | | | 0.4 | | | | 0.32 | | | |

Table shows estimated odds ratio (OR), 95% confidence interval (95%CI), p-value (*p*), with random-effects variances (var). Mixed effects logistic modelling estimating the association between the distribution of repellent and anti-gSG6-P1 IgG seropositivity, adjusting for temporality, season and risk group, with random-effects for village, month and village-specific heterogeneity in the effect of repellent.

*Indicates p-value for Wald joint tests for variables with >2 categories (risk group and season)

| Variable | 5 months use | | | | 6 months use | | | | 7 months use | | | |
|---|---|---|---|---|---|---|---|---|---|---|---|---|
| | **OR** | **95%CI** | | ***p*** | **OR** | **95%CI** | | ***p*** | **OR** | **95%CI** | | ***p*** |
| *Intervention* | | | | | | | | | | | | |
| No repellent | Ref. | | | | Ref. | | | | Ref. | | | |
| Repellent | 1.07 | 0.85 | 1.34 | 0.572 | 0.97 | 0.76 | 1.24 | 0.81 | 1.04 | 0.79 | 1.39 | 0.762 |
| *Risk group* | | | | *0.001 | | | | *0.001 | | | | *0.001 |
| Village Resident | Ref. | | | | Ref. | | | | Ref. | | | |
| Migrant | 0.86 | 0.73 | 1.02 | 0.081 | 0.86 | 0.73 | 1.02 | 0.083 | 0.85 | 0.71 | 1 | 0.052 |
| Forest Dweller | 1.21 | 1.08 | 1.36 | 0.001 | 1.22 | 1.09 | 1.37 | 0.001 | 1.21 | 1.08 | 1.36 | 0.001 |
| *Month* | 1.35 | 1.18 | 1.54 | 0 | 1.36 | 1.19 | 1.55 | 0 | 1.35 | 1.19 | 1.54 | 0 |
| *Season* | | | | *0.013 | | | | *0.013 | | | | *0.014 |
| Cool | Ref. | | | | Ref. | | | | Ref. | | | |
| Hot | 0.45 | 0.09 | 2.27 | 0.334 | 0.46 | 0.09 | 2.29 | 0.342 | 0.46 | 0.09 | 2.3 | 0.343 |
| Rainy | 0.14 | 0.04 | 0.54 | 0.004 | 0.14 | 0.04 | 0.54 | 0.004 | 0.14 | 0.04 | 0.55 | 0.004 |
| **Random effect variances** | **var** | | | | **var** | | | | **var** | | | |
| *Intercept: Month* | 1.15 | | | | 1.15 | | | | 1.15 | | | |
| *Intercept: Village* | 0.39 | | | | 0.38 | | | | 0.39 | | | |
| *Slope: Intervention* | 0.29 | | | | 0.35 | | | | 0.47 | | | |

Table shows estimated odds ratio (OR), 95% confidence interval (95%CI), p-value (*p*), with random-effects variances. Mixed effects logistic modelling estimating the association between the distribution of repellent and anti-gSG6-P1 IgG seropositivity, adjusting for temporality, season and risk group, with random-effects for village, month and village-specific heterogeneity in the effect of repellent.

*Indicates p-value for Wald joint tests for variables with >2 categories (risk group and season)

**Supplementary Table 5. Cumulative effects of repellent use on gSG6-P1 IgG antibody levels, modified by risk group (n=14,128 antibody measurements from N=10,857 participants).**

| Variable | 1 month use | | | | 2 months use | | | | 3 months use | | | | 4 months use | | | |
|---|---|---|---|---|---|---|---|---|---|---|---|---|---|---|---|---|
| | **MD** | **95%CI** | | ***p*** | **MD** | **95%CI** | | ***p*** | **MD** | **95%CI** | | ***p*** | **MD** | **95%CI** | | ***p*** |
| *Intervention* | | | | | | | | | | | | | | | | |
| No repellent | Ref. | | | | Ref. | | | | Ref. | | | | Ref. | | | |
| Repellent | 0.05 | 0.01 | 0.09 | 0.024 | 0.05 | 0.01 | 0.09 | 0.02 | 0.03 | -0.01 | 0.08 | 0.12 | 0.03 | -0.01 | 0.08 | 0.154 |
| *Risk group* | | | | *<0.001 | | | | *<0.001 | | | | *<0.001 | | | | *<0.001 |
| Village Resident | Ref. | | | | Ref. | | | | Ref. | | | | Ref. | | | |
| Migrant | 0 | -0.03 | 0.04 | 0.798 | 0 | -0.03 | 0.03 | 0.897 | 0 | -0.03 | 0.03 | 0.955 | -0.01 | -0.04 | 0.02 | 0.676 |
| Forest Dweller | 0.07 | 0.05 | 0.1 | <0.001 | 0.07 | 0.05 | 0.09 | <0.001 | 0.07 | 0.05 | 0.09 | <0.001 | 0.07 | 0.05 | 0.09 | <0.001 |
| *Intervention X Risk group* | | | | *0.010 | | | | *0.006 | | | | *0.003 | | | | *0.007 |
| Village Resident | Ref. | | | | Ref. | | | | Ref. | | | | Ref. | | | |
| Migrant | -0.06 | -0.12 | 0 | 0.041 | -0.08 | -0.14 | -0.02 | 0.014 | -0.09 | -0.16 | -0.03 | 0.005 | -0.07 | -0.14 | 0.01 | 0.075 |
| Forest Dweller | -0.05 | -0.08 | -0.01 | 0.007 | -0.05 | -0.08 | -0.01 | 0.007 | -0.05 | -0.09 | -0.01 | 0.008 | -0.06 | -0.1 | -0.02 | 0.003 |
| *Month* | 0.06 | 0.03 | 0.09 | <0.001 | 0.06 | 0.03 | 0.09 | <0.001 | 0.06 | 0.03 | 0.09 | <0.001 | 0.06 | 0.03 | 0.09 | <0.001 |
| *Season* | | | | *0.006 | | | | *0.005 | | | | *0.005 | | | | *0.005 |
| Cool | Ref. | | | | Ref. | | | | Ref. | | | | Ref. | | | |
| Hot | -0.15 | -0.51 | 0.22 | 0.435 | -0.14 | -0.51 | 0.22 | 0.436 | -0.14 | -0.51 | 0.22 | 0.44 | -0.14 | -0.51 | 0.22 | 0.434 |
| Rainy | -0.47 | -0.77 | -0.17 | 0.002 | -0.47 | -0.77 | -0.17 | 0.002 | -0.47 | -0.77 | -0.17 | 0.002 | -0.47 | -0.77 | -0.17 | 0.002 |
| **Random effect variances** | **var** | | | | **var** | | | | **var** | | | | **var** | | | |
| *Intercept: Month* | 0.06 | | | | 0.06 | | | | 0.06 | | | | 0.06 | | | |
| *Intercept: Village* | 0.02 | | | | 0.02 | | | | 0.02 | | | | 0.02 | | | |
| *Slope: Intervention* | 0.01 | | | | 0.02 | | | | 0.02 | | | | 0.02 | | | |

Table shows estimated mean difference (MD), 95% confidence interval (95%CI), p-value (*p*), with random-effects variances. Mixed effects linear modelling estimating the association between the distribution of repellent and anti-gSG6-P1 IgG levels, adjusting for temporality, season and the modifying effect of risk group (fitted as an interaction between intervention and risk group), with random-effects for village, month and village-specific heterogeneity in the effect of repellent.

*Indicates p-value for Wald joint tests for variables with >2 categories (risk group and season)

| Variable | 5 months use | | | | 6 months use | | | | 7 months use | | | |
|---|---|---|---|---|---|---|---|---|---|---|---|---|
| | **MD** | **95%CI** | | ***p*** | **MD** | **95%CI** | | ***p*** | **MD** | **95%CI** | | ***p*** |
| *Intervention* | | | | | | | | | | | | |
| No repellent | Ref. | | | | Ref. | | | | Ref. | | | |
| Repellent | 0.03 | -0.02 | 0.08 | 0.194 | 0.02 | -0.04 | 0.08 | 0.489 | 0.01 | -0.05 | 0.08 | 0.675 |
| *Risk group* | | | | *<0.001 | | | | *<0.001 | | | | *<0.001 |
| Village Resident | Ref. | | | | Ref. | | | | Ref. | | | |
| Migrant | -0.02 | -0.05 | 0.01 | 0.256 | -0.01 | -0.04 | 0.02 | 0.613 | -0.01 | -0.04 | 0.01 | 0.3 |
| Forest Dweller | 0.06 | 0.04 | 0.08 | <0.001 | 0.06 | 0.04 | 0.08 | <0.001 | 0.06 | 0.04 | 0.08 | <0.001 |
| *Intervention X Risk group* | | | | *0.008 | | | | *0.002 | | | | *0.004 |
| Village Resident | Ref. | | | | Ref. | | | | Ref. | | | |
| Migrant | 0.01 | -0.07 | 0.09 | 0.877 | -0.12 | -0.22 | -0.03 | 0.013 | -0.11 | -0.22 | 0 | 0.047 |
| Forest Dweller | -0.06 | -0.1 | -0.02 | 0.003 | -0.07 | -0.11 | -0.03 | 0.002 | -0.07 | -0.12 | -0.03 | 0.003 |
| *Month* | 0.06 | 0.03 | 0.09 | <0.001 | 0.07 | 0.04 | 0.09 | <0.001 | 0.07 | 0.04 | 0.09 | <0.001 |
| *Season* | | | | *0.005 | | | | *0.005 | | | | *0.005 |
| Cool | Ref. | | | | Ref. | | | | Ref. | | | |
| Hot | -0.15 | -0.51 | 0.22 | 0.426 | -0.14 | -0.51 | 0.22 | 0.435 | -0.14 | -0.51 | 0.22 | 0.442 |
| Rainy | -0.48 | -0.78 | -0.18 | 0.002 | -0.47 | -0.77 | -0.18 | 0.002 | -0.47 | -0.77 | -0.17 | 0.002 |
| **Random effect variances** | **var** | | | | **var** | | | | **var** | | | |
| *Intercept: Month* | 0.06 | | | | 0.06 | | | | 0.06 | | | |
| *Intercept: Village* | 0.02 | | | | 0.02 | | | | 0.02 | | | |
| *Slope: Intervention* | 0.02 | | | | 0.02 | | | | 0.03 | | | |

Table shows estimated mean difference (MD), 95% confidence interval (95%CI), p-value (*p*), with random-effects variances. Mixed effects linear modelling estimating the association between the distribution of repellent and anti-gSG6-P1 IgG levels, adjusting for temporality, season and the modifying effect of risk group (fitted as an interaction between intervention and risk group), with random-effects for village, month and village-specific heterogeneity in the effect of repellent.

*Indicates p-value for Wald joint tests for variables with >2 categories (risk group and season)

**Supplementary Table 6. Cumulative effects of repellent use on gSG6-P1 IgG antibody seropositivity, moderated by risk group (n=14,128 antibody measurements from N=10,857 participants).**

| Variable | 1 month use | | | | 2 months use | | | | 3 months use | | | | 4 months use | | | |
|---|---|---|---|---|---|---|---|---|---|---|---|---|---|---|---|---|
| | **OR** | **95%CI** | | ***p*** | **OR** | **95%CI** | | ***p*** | **OR** | **95%CI** | | ***p*** | **OR** | **95%CI** | | ***p*** |
| *Intervention* | | | | | | | | | | | | | | | | |
| No repellent | Ref. | | | | Ref. | | | | Ref. | | | | Ref. | | | |
| Repellent | 1.23 | 0.98 | 1.54 | 0.077 | 1.16 | 0.91 | 1.47 | 0.222 | 1.06 | 0.82 | 1.37 | 0.675 | 1.19 | 0.91 | 1.55 | 0.197 |
| *Risk group* | | | | *<0.002 | | | | *<0.003 | | | | *<0.004 | | | | *<0.005 |
| Village Resident | Ref. | | | | Ref. | | | | Ref. | | | | Ref. | | | |
| Migrant | 0.83 | 0.68 | 1.03 | 0.088 | 0.83 | 0.68 | 1.02 | 0.071 | 0.85 | 0.7 | 1.03 | 0.096 | 0.85 | 0.71 | 1.02 | 0.085 |
| Forest Dweller | 1.29 | 1.11 | 1.51 | 0.001 | 1.3 | 1.12 | 1.5 | <0.001 | 1.3 | 1.13 | 1.5 | <0.001 | 1.3 | 1.13 | 1.48 | <0.001 |
| *Intervention X Risk group* | | | | *0.263 | | | | *0.218 | | | | *0.298 | | | | *0.074 |
| Village Resident | Ref. | | | | Ref. | | | | Ref. | | | | Ref. | | | |
| Migrant | 1.13 | 0.79 | 1.6 | 0.512 | 1.11 | 0.76 | 1.61 | 0.589 | 0.99 | 0.66 | 1.48 | 0.957 | 0.98 | 0.62 | 1.53 | 0.917 |
| Forest Dweller | 0.87 | 0.69 | 1.09 | 0.217 | 0.85 | 0.67 | 1.06 | 0.152 | 0.83 | 0.66 | 1.06 | 0.133 | 0.75 | 0.58 | 0.97 | 0.027 |
| *Month* | 1.34 | 1.18 | 1.53 | <0.001 | 1.35 | 1.18 | 1.54 | <0.001 | 1.36 | 1.2 | 1.55 | <0.001 | 1.36 | 1.19 | 1.54 | <0.001 |
| *Season* | | | | *0.015 | | | | *0.014 | | | | *0.016 | | | | *0.015 |
| Cool | Ref. | | | | Ref. | | | | Ref. | | | | Ref. | | | |
| Hot | 0.46 | 0.09 | 2.33 | 0.347 | 0.46 | 0.09 | 2.32 | 0.349 | 0.48 | 0.1 | 2.4 | 0.371 | 0.46 | 0.09 | 2.33 | 0.351 |
| Rainy | 0.14 | 0.04 | 0.55 | 0.005 | 0.15 | 0.04 | 0.55 | 0.005 | 0.15 | 0.04 | 0.57 | 0.005 | 0.15 | 0.04 | 0.56 | 0.005 |
| **Random effect variances** | **var** | | | | **var** | | | | **var** | | | | **var** | | | |
| *Intercept: Month* | 1.17 | | | | 1.15 | | | | 1.15 | | | | 1.14 | | | |
| *Intercept: Village* | 0.37 | | | | 0.36 | | | | 0.35 | | | | 0.37 | | | |
| *Slope: Intervention* | 0.22 | | | | 0.3 | | | | 0.41 | | | | 0.34 | | | |

Table shows estimated odds ratio (OR), 95% confidence interval (95%CI), p-value (*p*), with random-effects variances. Mixed effects logistic modelling estimating the association between the distribution of repellent and anti-gSG6-P1 IgG seropositivity, adjusting for temporality, season and the modifying effect of risk group (fitted as an interaction between intervention and risk group), with random-effects for village, month and village-specific heterogeneity in the effect of repellent.

*Indicates p-value for Wald joint tests for variables with >2 categories (risk group and season)

| Variable | 5 months use | | | | 6 months use | | | | 7 months use | | | |
|---|---|---|---|---|---|---|---|---|---|---|---|---|
| | **OR** | **95%CI** | | ***p*** | **OR** | **95%CI** | | ***p*** | **OR** | **95%CI** | | ***p*** |
| *Intervention* | | | | | | | | | | | | |
| No repellent | Ref. | | | | Ref. | | | | Ref. | | | |
| Repellent | 1.24 | 0.95 | 1.63 | 0.118 | 1.25 | 0.93 | 1.67 | 0.141 | 1.41 | 1 | 1.99 | 0.048 |
| *Risk group* | | | | *<0.006 | | | | *<0.007 | | | | *<0.008 |
| Village Resident | Ref. | | | | Ref. | | | | Ref. | | | |
| Migrant | 0.86 | 0.72 | 1.03 | 0.104 | 0.91 | 0.76 | 1.08 | 0.27 | 0.88 | 0.74 | 1.05 | 0.145 |
| Forest Dweller | 1.31 | 1.15 | 1.49 | <0.001 | 1.33 | 1.17 | 1.51 | <0.001 | 1.3 | 1.15 | 1.48 | <0.001 |
| *Intervention X Risk group* | | | | *0.034 | | | | *0.007 | | | | *0.007 |
| Village Resident | Ref. | | | | Ref. | | | | Ref. | | | |
| Migrant | 1.08 | 0.65 | 1.81 | 0.756 | 0.67 | 0.38 | 1.18 | 0.166 | 0.66 | 0.33 | 1.32 | 0.24 |
| Forest Dweller | 0.72 | 0.55 | 0.94 | 0.017 | 0.64 | 0.48 | 0.85 | 0.002 | 0.6 | 0.43 | 0.82 | 0.002 |
| *Month* | 1.35 | 1.18 | 1.54 | <0.001 | 1.36 | 1.19 | 1.55 | <0.001 | 1.36 | 1.19 | 1.54 | <0.001 |
| *Season* | | | | *0.014 | | | | *0.014 | | | | *0.014 |
| Cool | Ref. | | | | Ref. | | | | Ref. | | | |
| Hot | 0.45 | 0.09 | 2.28 | 0.335 | 0.46 | 0.09 | 2.3 | 0.344 | 0.46 | 0.09 | 2.31 | 0.344 |
| Rainy | 0.14 | 0.04 | 0.54 | 0.004 | 0.14 | 0.04 | 0.54 | 0.004 | 0.14 | 0.04 | 0.55 | 0.005 |
| **Random effect variances** | **var** | | | | **var** | | | | **var** | | | |
| *Intercept: Month* | 1.15 | | | | 1.15 | | | | 1.15 | | | |
| *Intercept: Village* | 0.38 | | | | 0.37 | | | | 0.39 | | | |
| *Slope: Intervention* | 0.29 | | | | 0.33 | | | | 0.45 | | | |

Table shows estimated odds ratio (OR), 95% confidence interval (95%CI), p-value (*p*), with random-effects variances. Mixed effects logistic modelling estimating the association between the distribution of repellent and anti-gSG6-P1 IgG seropositivity, adjusting for temporality, season and the modifying effect of risk group (fitted as an interaction between intervention and risk group), with random-effects for village, month and village-specific heterogeneity in the effect of repellent.

*Indicates p-value for Wald joint tests for variables with >2 categories (risk group and season)

Instantaneous

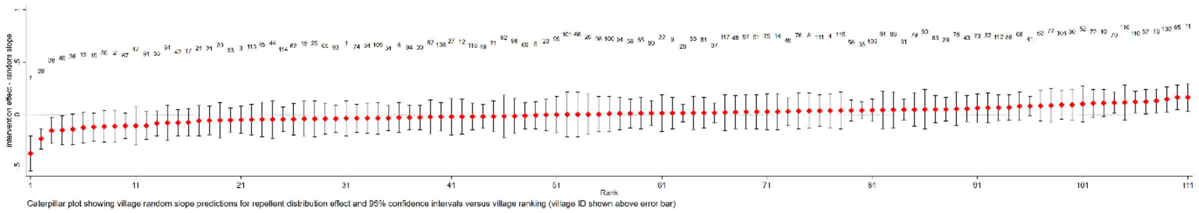


3 months use

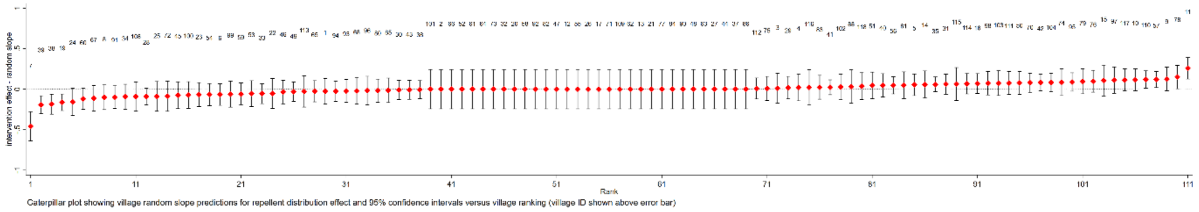


6 months use

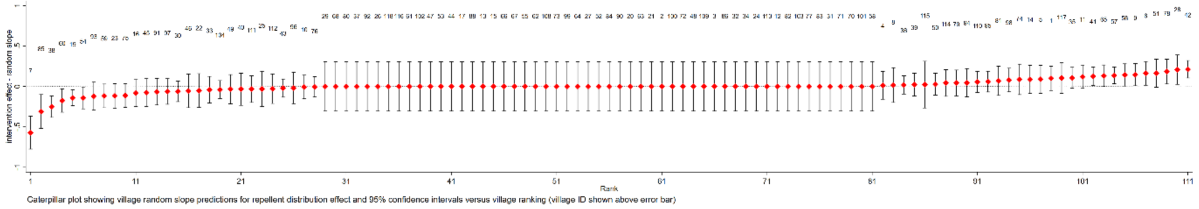


**Supplementary Figure 4. Caterpillar plot showing village random slope predictions for effect of repellent use (instantaneous and cumulative (3 and 6 months)) on anti-gSG6-P1 IgG levels and 95% confidence intervals for 114 villages sorted by estimated mean difference value.**

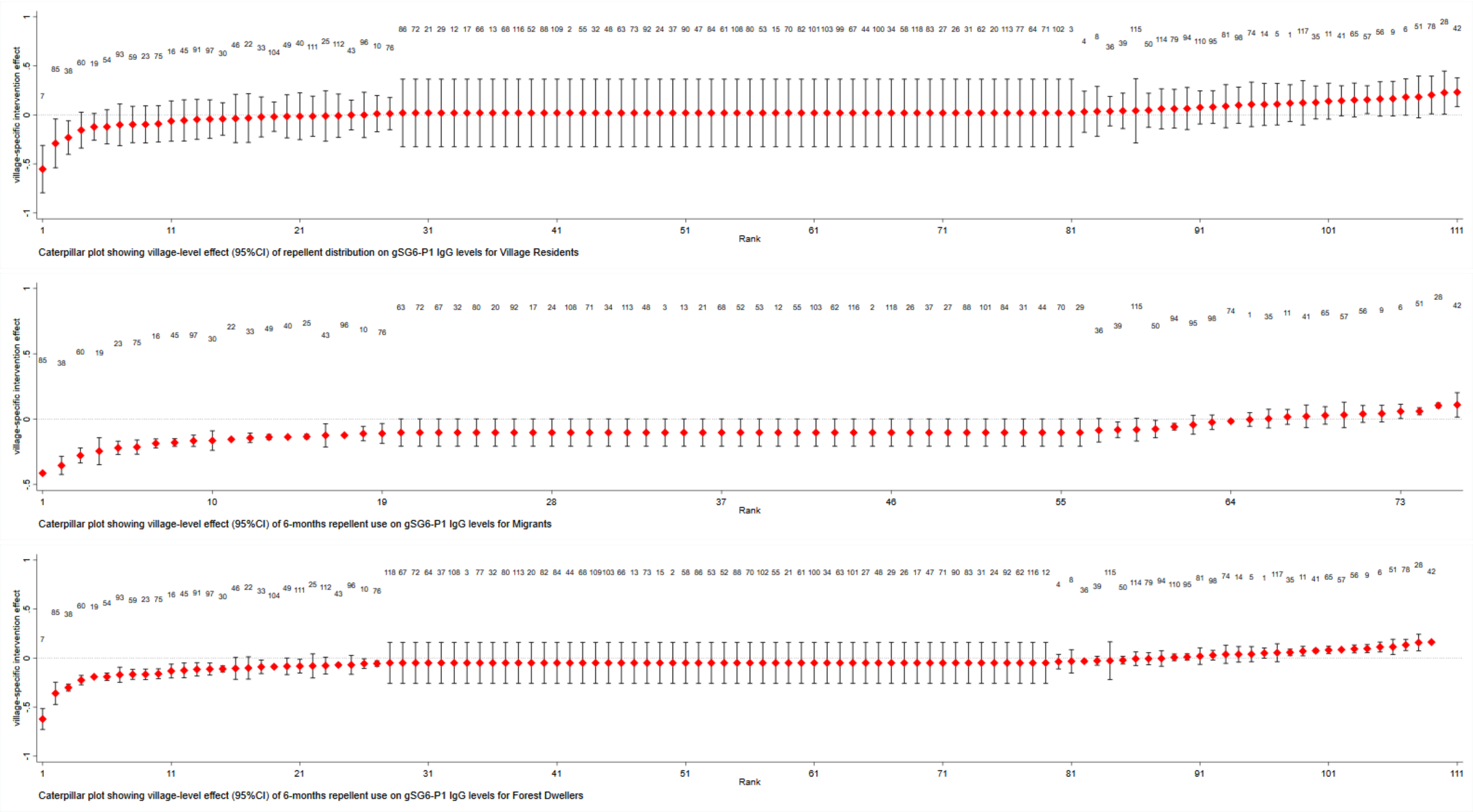


**Supplementary Figure 5. Caterpillar plots show village-level effect with 95% confidence intervals of 6-months of repellent use on gSG6-P1 IgG levels for each risk group.** Estimates were derived by combining village-specific random effects (*i.e.* slope) with the fixed effects estimates of 6-months use the intervention on gSG6-P1 IgG levels for village residents, migrants and forest dwellers.
